# Vagina reconstruction by decellularization of healthy human vaginal tissue

**DOI:** 10.1101/2023.04.05.23288175

**Authors:** J. Sueters, F. Xiao, J.P.R.W. Roovers, M.B. Bouman, F.A. Groenman, H. Maas, J.A.F. Huirne, T.H. Smit

## Abstract

When a healthy, full-size vagina is absent due to a disorder, various neovagina creation methods are available. Sometimes dilation or stretching of the vagina cavity is sufficient, but generally intestinal or dermal graft tissue is required. However, different inherent tissue properties cause complications. Therefore, when a body part is lost, it should be replaced by a similar material. The use of organ-specific acellular vaginal tissue carries great potential, as the similar architecture and matrix composition make it fit for vagina regeneration. We developed an optimized decellularization protocol for human vaginal tissue and determined suitability as tissue-mimicking scaffold for vagina reconstruction. Histological examination confirmed the preservation of structural features and minimal cellular residue was seen during fluorescence microscopy, DNA and RNA quantification and fragment-length examination. Biomechanical testing showed decreased (P<0,05) strain at rupture (23%), tensile stress (55%) and elastic modulus (68%) after decellularization. Fluorescence microscopy revealed preserved Fibronectin-I/II/III and Laminin-I/II, while Collagen-I and Ficolin-2B were decreased but mostly retained. The absence of cellular residue, minimally altered biomechanical ECM properties and mostly preserved structural proteins, appear to make our decellularized human vaginal matrix a suitable tissue-mimicking scaffold for vagina transplantation when tissue survival through vascularization and innervation are accomplished in the future.

## Introduction

Absence of a functional vagina is caused by various medical disorders of congenital (Mayer-Rokitansky-Küster-Hauser syndrome, cloacal malformations, endocrine abnormalities, gender dysphoria and disorder of sex development) or acquired origin (like cancer and trauma), that tremendously reduce the quality of life (QoL) and psychological well being [1]. This often requires partial or total reconstruction, aiming to construct a vagina with function and sensation similar to that of a native vagina. Vaginoplasty is mostly performed on patients with Mayer-Rokitansky-Küster-Hauser syndrome (MRKHS) or Male-to-Female Gender Dysphoria (MtF GD). MRKHS presents as congenital aplasia of uterus and upper two-third of the vagina with normal secondary sexual characteristics [2], with approximately 39.000-650.000^1^ annual vaginoplasties. Non-surgical alternatives (see [3]–[7]) carry great disadvantages [8], [9] and regularly require secondary surgical treatment [10]. Expression of GD, an incongruency between assigned sex at birth and gender, is diverse. Gender assignment surgery (GAS) is not always desired, but roughly 397.000^2^ annual vaginoplasties are performed to increase patient QoL and sexual function [11]–[14]. Additional grafts are regularly required due to insufficient local, autologous skin volume due to puberty hormone blockers at early age [15]–[19].

For vaginoplasty over 20 grafting approaches with characteristic (dis)advantages have been developed without golden standard [20], [21]. Allogenic- and xenografts pose high risk of immunorejection [21], [22], xenografts also carry cross-specie transmission risk of infections and diseases [23], [24]. Therefore, vaginoplasty generally relies on autologous dermal or intestinal grafts [25], [26], but heterotopic vagina-mimicking grafts are inherently different in physiology [27]. Consequent lack of or excessive vaginal mucus production, aesthetic issues and inadequate neovaginal lengths regularly requires revisional surgery [28]–[33]. Hence, vaginoplasty outcome is not always satisfactory, with substantial room for improvement [11], [29], [32], [34]–[36].

Tissue engineering offers a great alternative, as it can theoretically create any tissue or organ through infinite combinations of three essentials: 1) cells (type and state), 2) biomaterial (composition, mechanical properties, texture and condition) and 3) signaling factors (e.g., growth factors and bioreactors) [37]. Tissue-engineered organs have found successful clinical applications [38], [39], with decellularized cartilage [40], [41], bone [42], tendon [43]–[45], heart [46]–[49], lung [50], [51], vessel [52]–[61], skeletal muscle [62], [63], intestine [64], liver [65]–[68], pancreas [69], [70], kidney [71]–[74], bladder [75], [76], cornea [77]–[82] and uterus [83]. This field is challenging due to infinite tunability, but current *in vitro* cultured neovaginal constructs offer great promise for autogenic grafts without large donor site harvesting and associated risks [84]–[88].

We previously reviewed used tissue-engineering applications for vaginoplasty [37]. Most included articles only reported *in vitro* experiments [84]–[88] or small transplants in rodents [56], [89], [98], [99], [90]–[97]. In clinically relevant sizes (centimetre-range and larger), tissue survival is restricted by diffusion and mass transport of nutrients [100]. Mass transport is also crucial directly after implantation, as a vascular network is not established yet. Constructs of clinically relevant sizes mostly incorporated Extracellular Matrix (ECM) biomaterials, reconstructed partial vaginas [38], [89], [101]–[106] and applied cell-free grafts [102]–[111]. Clinical trials reported granulation, bacterial infections and inflammation in cell-free scaffolds [99], [105]–[109], whereas autologous cells reduced fibrosis and increased regeneration and satisfactional, complication-free results [112]– [114].

In theory, vaginal matrices form the ideal biomaterial, to prevent complications from inherent physiological differences. An Acellular Vaginal Matrix (AVM) from rats [94], [97] or pigs [85], [99] has been tested *in vitro* [85] and transplanted in rats [94], [97], [99]. Partial, cell-free AVM constructs resulted in adhesion without chronic inflammation or fibrosis and good biomechanical properties, biocompatibility and presence of various growth factors [97], [99]. Despite successful isolation of human Vaginal Epithelial Cells, Vaginal Stromal Cells, Vaginal Fibroblasts and Vaginal Mucosa [95], [96], [115], [116], human AVM has only been created once from Pelvic Organ Prolapse-patients [117].

In our medical centre, colpectomy was performed 60 times in 2022 in transgender men, due to feelings of gender dysphoria, to reduce vaginal discharge or to prevent fistula formation in urethral lengthening for phallo- or metaidoioplasty [118]. Our aim was to use colpectomy waste tissue to create human decellularized Vaginal Matrix. Remnant cellular material within ECM may cause cytocompatibility issues and adverse host responses *in vivo* [119], [120] above (unknown) threshold concentrations. Based on *in vivo* remodeling-reports, the following criteria suffice to assess successful decellularization: (1) No nuclear material visible, (2) <50 ng OR <10% dsDNA/mg ECM dry weight and (3) DNA fragments <200 base pair [121]. To assess functionality, three criteria were added. Avoidance of chemically induced structural damage, was defined as: (4) visible structural features. Furthermore, elasticity is a predominant feature of the vagina that allows elongation during intercourse and passage of a full-term baby during birth, this was investigated by: (5) biocompatibility during stretching. Lastly, vagina characteristics greatly depend on interaction between cells, extracellular matrix (ECM) and its various structural proteins, thus biocompatibility was further investigated by: (6) presence of visible collagen, elastin, laminin and fibronectin.

## Materials & Method

### Patients and surgical procedure

Vaginal tissue was retrieved from 12 patients during robotic-assisted laparoscopic colpectomy at Amsterdam UMC location VU Medical Center in Amsterdam between April 2022 and December 2022. All procedures were done using the daVinci XI system (Intuitive, Madrid, Spain). First, vaginal epithelium was carefully dissected with monopolar scissors and fenestrated bipolar forceps to prevent bleeding as much as possible [122]. Dissection was performed approximately 2 centimeters proximal of the ostium urethrae externum (marked by a suture) and up to the posterior commissure level [122]. Thin layer and not full thickness vaginal tissue was removed, to prevent nerve injury to adjacent structures, fistula to bladder, urethra or rectum and bleeding from the perivaginal plexus [122]. Per patient, two proximal rings (Mesoderm) originating from paramesonephric duct and a distal ring (Ectoderm) from urogenital sinus were obtained. The vaginal apex was sutured laparoscopically by suturing endopelvic fascia of the vesicovaginal space and remnants of rectovaginal septum together [122]. If necessary, residual introital epithelium was removed vaginally and the introitus was narrowed through approximation of bulbocavernosus muscles with 1-2 sutures [122]. Vaginal tissue was directly stored in phosphate buffered saline (PBS) with pH 7,4 on ice until decellularization. Written informed consent for experimental use of vaginal waste tissue was obtained from all patients. Tissue collection and experimental usage was approved by the institutional Medical Ethical Examination Committee of Amsterdam UMC location VU Medical Center (Amsterdam; IRB approval by METc VUmc registration number 2018/3190, October 2018).

### Decellularization

Decellularization is commonly based on chemical, biological and/or physical processes, depending on tissue origin, size and type [121], [123]. Our DC protocol excluded physical processes to prevent ECM disruption [121], [123] and was derived from previous protocols for whole, large organ vaginal tissue [85], [99], [117]. We applied Triton X-100, Sodium deoxycholate and DNase I [85], [117] at minimally required concentrations, to prevent diminished growth factors, proteins, mechanical properties and ultrastructure [121], [123]. Detrimental effects on ECM were minimized by keeping maximum exposure time to reagent under 24 hours [121]. Decellularization was initiated within 4 h by 24 h incubation at 37°C with constant agitation (100 motions/minute) in 0,18% w/w Triton x-100 (Sigma-Aldrich, st. Louis, MO, USA) and 0,05% w/w sodium-deoxycholate (Sigma-Aldrich, st. Louis, MO, USA) in PBS (Fresenius Kabi, Zeist, Utrecht, the Netherlands) [124]. Tissue was washed 20 min. in PBS twice and washed 72 h at 4°C with constant agitation (on a roller) in PBS with 1% penicillin/ streptomycin (100 U/mL penicillin and 100 μg/mL streptomycin; Life Technologies Europe BV, Bleiswijk, Zuid-Holland, the Netherlands). Enzymatic digestion involved 24 h incubation at 37°C for by DNase I (150 IU/mL; Sigma-Aldrich, st. Louis, MO, USA) and 50 mmol MgCl_2_ (Sigma-Aldrich, st. Louis, MO, USA) in PBS. Before further processing, 24 h incubation at 4°C with constant agitation in 1% penicillin/streptomycin PBS and an extensive PBS wash were performed. The total protocol duration was 6 days.

### DAPI with Ethidium Bromide or Haematoxylin with Eosin Staining

DAPI and EtBr bind to nuclear cellular material with distinctive binding sensitivity and fluorescence intensity due to inherent differences in binding method.^3^ DAPI- and EtBr-staining combined allows extensive assessment of nuclear cellular material with Blue/Purple-stained dsDNA and Red-stained RNA [125], [126]. A 4’,6-diamidino-2-phenylindole (DAPI) with Ethidium Bromide (EtBr) staining and Haematoxylin with Eosin (H&E) staining was performed on native and decellularized vaginal tissue. Samples were fixed overnight at RT with constant agitation (on a roller) in 4% w/v formaldehyde (ROTI® Histofix 4%; Carl Roth, Karlsruhe, Mannheim, Germany). Tissue was washed 30 min. in PBS and thrice for 30 min. in 70% ethanol (Sigma-Aldrich, st. Louis, MO, USA) at RT. Tissue was embedded according to *Embedding by semi-automated tissue processing* (below). Samples were sliced with a Leica RM2255 microtome (Leica Biosystems, Deer Park, IL, USA) with a 6° knife angle to 5 μm-thick sections and sections were mounted on microscope slides with the Leica HI1210 water bath (Leica Biosystems, Deer Park, IL, USA) at 37,2°C. The slide-sections are deparaffinized 5 min. in xylene (twice) and hydrated by 2 min. sequential steps in 100% ethanol (twice), 96% ethanol and 70% ethanol. Staining involved 5 min. incubation at RT in either 0,2 μg/ml DAPI (Sigma-Aldrich, st. Louis, MO, USA) in PBS or 5 min. incubation in Haematoxylin (Sigma-Aldrich, st. Louis, MO, USA) and 3 min. in Eosin (Sigma-Aldrich, st. Louis, MO, USA). H&E-stained slides were embedded with Entellan (Sigma-Aldrich, st. Louis, MO, USA) and dried overnight before examination with the Leica DM5000B (Leica Biosystems, Deer Park, IL, USA) fluorescence microscope. DAPI-stained slides were embedded with Prolong® Gold Antifade Reagent (Life Technologies Europe BV, Bleiswijk, Zuid-Holland, the Netherlands), hardened 1 h at RT and placed at 4°C for several hours before analysis with the Olympus BX 41 (Olympus, Tokyo, Japan) light microscope.

### Embedding by semi-automated tissue processing

Tissues were embedded in the Epredia™ Excelsior™ AS Tissue Processor (Thermo Fisher Scientific, Landsmeer, Zuid-Holland, the Netherlands). This sequentially involved 1 h in 70% ethanol, 1 h in 90% ethanol, 1 h in 96% ethanol and (thrice) 1 h in 100% ethanol at RT. Three sequential xylene steps at 37°C, 40°C and 45°C were performed, followed by sequential paraffin bath 1, 2 and 3 in three steps of each 1 h and 20 min at 62°C. Next, tissue is manually embedded in liquid paraffin of 55°C by 1 h solidification at RT on a cooled plate of -5°C. After overnight 4°C paraffin wax hardening, samples are ready for microtome sectioning.

### Quantitative DNA and RNA analysis

DNA and RNA residues were isolated with the commercial DNeasy Blood & Tissue kit (Qiangen, Shenzhen, Nanshan, China). In brief, a maximum of 25 mg of thawed native or wet DC vaginal tissue were placed in microcentrifuge tubes with 180 μL buffer ATL and 20 μL proteinase K. After vortexing, the tubes were placed on a 56°C heated tube-holder plate until complete digestion (approximately 90 min.) with 15 min. interval vortexing. Lysis buffer AL in ethanol was added and after 1 min. centrifugation at 6.000 x G, several washing steps were performed to remove contaminates. The residue mixture was diluted by buffer AE. DNA concentration was measured by the Qubit 2.0 fluorometer (Thermo Fisher Scientific, Landsmeer, Zuid-Holland, the Netherlands) with the dsDNA HS Assay kit (Thermo Fisher Scientific, Landsmeer, Zuid-Holland, the Netherlands) and optimized working solution - DNA sample ratio for the 0,1-120 ng/μL application range. RNA concentration was measured by the Qubit 2.0 fluorometer with the RNA HS Assay kit (Thermo Fisher Scientific, Landsmeer, Zuid-Holland, the Netherlands) and optimized working solution - RNA sample ratio for the 4-200 ng/μL application range. The RNA and DNA quantity per mg of dry ECM tissue was calculated.

### DNA content analysis

DNA concentration in DC vaginal tissue was too low to assess DNA fragments by gel electrophoresis. After quantitative DNA analysis, DNA was diluted in PBS to 1 ng/μL and analysed in the Agilent 4200 TapeStation System (Agilent Technologies, Amstelveen, Noord-Holland, the Netherlands) to assess fragment length.

### Biomechanical analysis of tensile stress, strain at rupture and elastic modulus

Wet native and DC vaginal tissue were mechanically loaded using the EBERS TC-3 mechanical stimulation bioreactor (EBERS Medical Technology SL, Zaragoza, Spain). Samples were elongated along top-to-bottom direction. Section dimensions were measured with a vernier calliper and short strips of (3,4±0,5) x (3,2±1,2) mm [native] or (3,9±0,8) x (3,1±0,9) mm [DC] in width x thickness, were clamped between two flat grips with an initial 5-mm gap. With 120 mm/min. elongation speed, the test was stopped at point of full rupture. At least seven biological replicates were tested for each sample condition, with averaging over 10 consecutive load cycles and 1 mm addition elongation steps. Strain was calculated as the elongated displacement ratio to initial length ΔL/L_0_ [99], [127]. Ultimate tensile stress σ was calculated from peak load *F*, mean width *w* and thickness *t* as σ = *F*/(*w* x *t*) [99], [127]. By plotting σ (ΔL/L_0_), the strain at rupture ε was determined as well as the elastic modulus *E* = 0,4 x σ/(ε_60%σ_ - ε_20%σ_). Here, strain ε_60%σ_ and ε_20%σ_ corresponded to 60% and 20% of σ, respectively [99], [127].

Elastic behaviour of intact vaginal rings of (27,4±1,9) x (4,4±0,9) x (2,6±0,5) mm [native] or (26,1±3,5) x (3,8±0,8) x (2,9±0,5) mm [DC] in dimensions length *l* x *w* x *t*, was determined with a home-made setup. Non-dissolvable sutures were placed around both ends of the ring, one was clamped and the second was placed behind a hook. Mechanical loading of 10 consecutives cycles was performed by software driven electronic movement of the hook at 6 mm/min. for pre-determined strains. The peak load *F* was determined as previously described.

### Immunofluorescence analysis of ECM constitutive proteins

Immunofluorescence of constitutive ECM proteins was performed on native and DC vaginal tissue. Slides were deparaffinized and rehydrated according to *DAPI with Ethidium Bromide or Haematoxylin with Eosin staining* (above) and washed 5 min. in PBS (thrice) while slowly shaking. Antigen retrieval was performed 10 min. for Collagen-I and Ficolin-2B by 10 mM Tris (Roche, #10708976001, Woerden, Utrecht, the Netherlands) + 1 mM EDTA (Bio-rad, #161-0770, Lunteren, Gelderland, the Netherlands) at pH=9,0. Auto fluorescence was quenched for Laminin- and Fibronectin-protocols by 30 sec. incubation with TrueBlack® (Biotium, #23007, Fremont, CA, USA) and slides were washed 5 min. in PBS (thrice) while slowly shaking. A-specific binding of antibodies for Laminin and Fibronectin was minimized by 1 h incubation with Superblock™ (ThermoFischer Scientific, #37515, Landsmeer, Zuid-Holland, the Netherlands) and for Collagen and Ficolin by 1 h incubation with 3% BSA – IgG free (Sigma, #A0281-5G, st. Louis, MO, USA) + 0,1% Tween-20 (Sigma, #P1379, st. Louis, MO, USA) + 0,1% BSAc – IgG free (Sigma, st. Louis, MO, USA) in a humid slide box at RT. A short and gentle PBS rinse was performed. Slides were incubated with fluorescent conjugated antibodies for Fibronectin-I/II/III (Abcam, #ab198934, rabbit mAb, 1:100 concentration, overnight incubation at 4°C) and Laminin-I/II (ThermoFischer Scientific, #PA5-22901, rabbit pAb, 1:100 concentration, overnight incubation at 4°C) diluted in Bright Diluent (VWR, #UD09-500, Amsterdam, Noord-Holland, Amsterdam) or Collagen-I (Bioss antibodies, #BS-10423R-A350 and BS-0709R-A350, 1:50 concentration, overnight incubation at 4°C) and Elastin (Bioss antibodies, #BS-13162R-A594, 1:50 concentration, overnight incubation at 4°C) diluted in 3% BSA – IgG free (Sigma, #A0281-5G, st. Louis, MO, USA) + 0,1% Tween-20 (Sigma, #P1379, st. Louis, MO, USA). After 30 min. accumulation at RT, slides were washed 10 min. in PBS (thrice) while slowly shaking. Ficolin-Laminin- and Fibronectin-slides were incubated 5 min. with 300 ng/mL DAPI in PBS and Collagen-slides for 15 min. with 1 μg/ml EtBr (Applichem, #A1152.0010, Darmstadt, Germany) in PBS. Slides were washed 5 min. in PBS and embedded with Prolong® Gold Antifade Reagent (Life Technologies Europe BV, Bleiswijk, Zuid-Holland, the Netherlands), hardened 1 h at RT and placed at 4°C for a couple of hours before analysis with the Olympus BX 41 (Olympus, Tokyo, Japan) light microscope.

### Statistical analysis

For feasibility, vaginal tissue from 12 patients was obtained to perform experiments with 7 biological and 3 technical replicates per protocol. Statistical analysis was performed to identify significant differences between native and decellularized tissue, concerning DNA and RNA quantity and biomechanical properties. Results were expressed as mean with standard deviation (SD). As a normal distribution was absent and a difference between two individual conditions was assessed, the Mann Whitney U-test was applied. The P-value was obtained in a two-tailed test and was considered statistically significant for *P≤0,05 and **P≤0,01 and ***P≤0,001.

## Results

### Gross anatomic characteristics of DC vaginal scaffolds

Vaginal removal is a delicate procedure due to the thin wall layers, where also elasticity and coagulation are patient-dependent and affected procedure difficulty. Vaginal tissue could successfully be removed by dissection in multiple rings to prevent bleeding from the well-vascularized vaginal wall and to avoid damage to adjacent structures. Two mesoderm rings and an ectoderm ring (Figure 1A) or vaginal tissue with tears (Figure 1B) were removed per patient. Patient mean age was 27,1 (range 19-51) years with mean 5,9 (range 2-11) years presurgical androgen exposure by testosterone injection (Nebido [Bayer Healthcare], Sustanon [Aspen]) or testosterone gel (Androgel [Besins Healthcare]). Burned and cankered (i.e., ragged, lacerated or thinned) sections were removed, after which tissue was suitable for analysis. Native tissue was sectioned to <25 mg dry weight for storage compatibility and either fixed in 70% ethanol at 4°C for microscopic examination with DAPI- and H&E-staining or dried and snap-frozen in liquid N_2_ with -80°C storage for quantitative DNA- and RNA-analysis. Decellularized full vagina rings were sectioned for analysis if needed. Decellularization of all 12 patient samples was without issues with visually, a colour-change from pink native to white DC tissue was observed (Figure 1D), that initially indicates (some or) successful DC.

**Figure 1:**
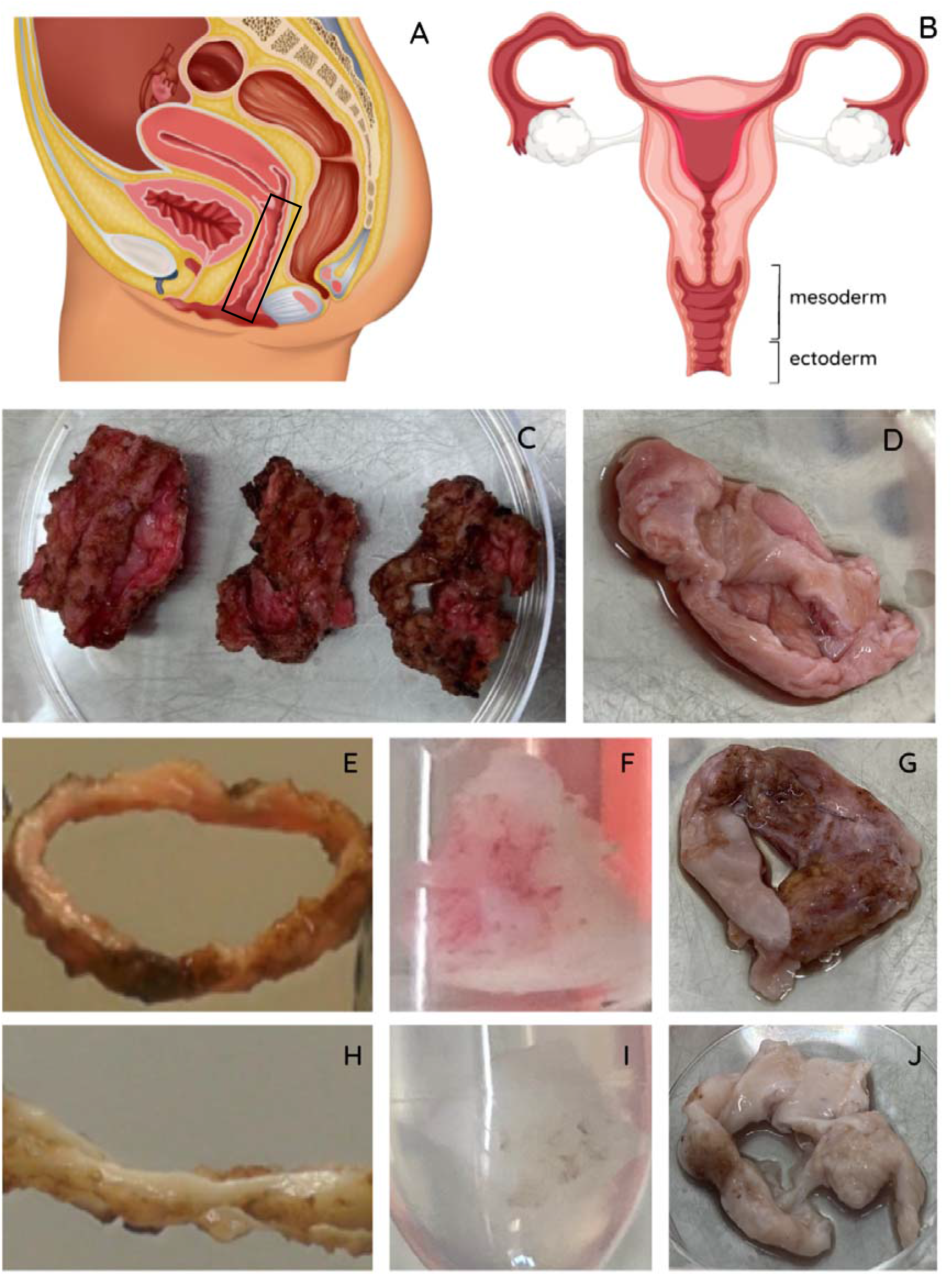
A) Illustration of female reproductive system sideview with B) fractions of mesoderm tissue (2/3 proximal vagina) and ectoderm tissue (1/3 distal vagina) front view C) Completely removed vaginal rings from a single patient during robot-assisted laparoscopic colpectomy. Orientation proximal-distal origin from left to right. Image was taken before removal of burned and cankered tissue prior to the decellularization protocol. D) Removed vaginal tissue from a single patient during vaginally performed colpectomy. Image was taken prior to the decellularization protocol. Macroscopic inspection of successful decellularization (DC) showed pink vagina before DC in E) ring, F) block and G) section, and white vaginal tissue after DC in H) ring, I) block and J) section.

### Structural features with no nuclear cellular material visible after DC

DAPI- with EtBr-staining confirmed absence of visible DNA and RNA after DC (Figure 2C), and abundant presence in positive controls of mesoderm (Figure 2A) and ectoderm (Figure 2B) sections. Decellularization by enzymatic digestion (Figure 2D) removed visible RNA and the majority of DNA successfully. After decellularization by membrane disruption, DNA and RNA remained visible (Figure 2E).

**Figure 2:**
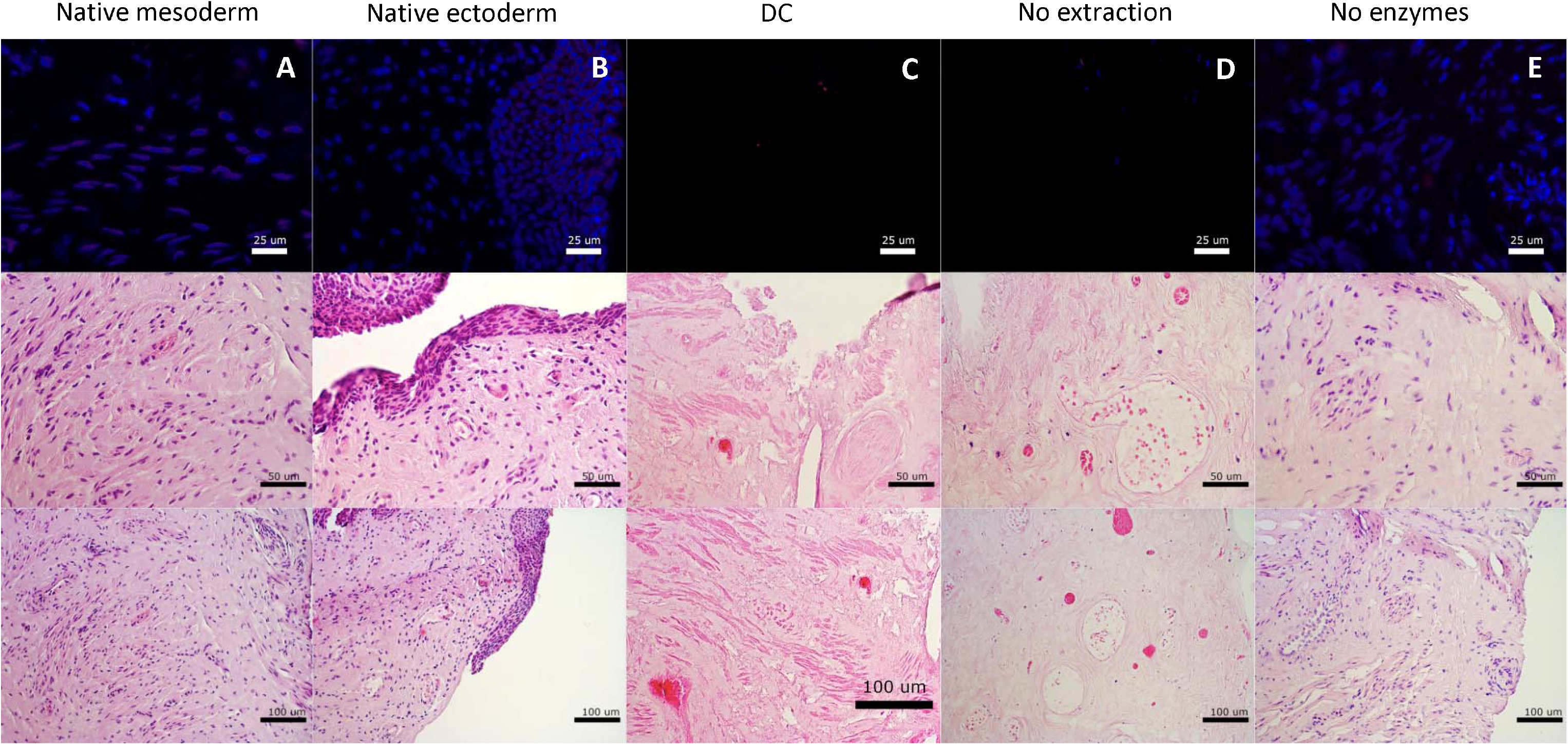
Histological assessment with DAPI and Ethidium Bromide (EtBr) staining (top) and assessment of tissue structure with Haematoxylin and Eosin (H&E) staining (center and bottom) of A) native mesoderm, B) native ectoderm vaginal tissue, C) decellularization, D) Enzymatic degradation only and E) Membrane disruption only.

H&E-staining confirmed intact structures and absence of visible nuclei (Figure 2C) after DC, that were abundantly present in native mesoderm (Figure 2A) and ectoderm (Figure 2B) sections. Decellularization by membrane disruption preserved structure and visible nuclei (Figure 2E). Decellularization by enzymatic degradation (Figure 2D) caused structural alterations.

Decellularization efficacy was further assessed on full size vagina wall rings for clinically relevant tissue sizes (Figure 3). H&E-staining confirmed absence of visible nuclear components and intact structures in decellularized mesoderm (Figure 3B) and ectoderm (Figure 3C) rings. Double chemical concentrations for decellularization (Figure 3D) resulted often in compromised structures and tissue became physically difficult to handle, due to extreme softening. Vaginal tissue was not compromised by (long-term) presurgical androgen exposure. No epithelial atrophy (thinned squamous epithelium) or prostatic metaplasia (glandular structures in epithelium or subepithelial stroma) were assessed.

**Figure 3:**
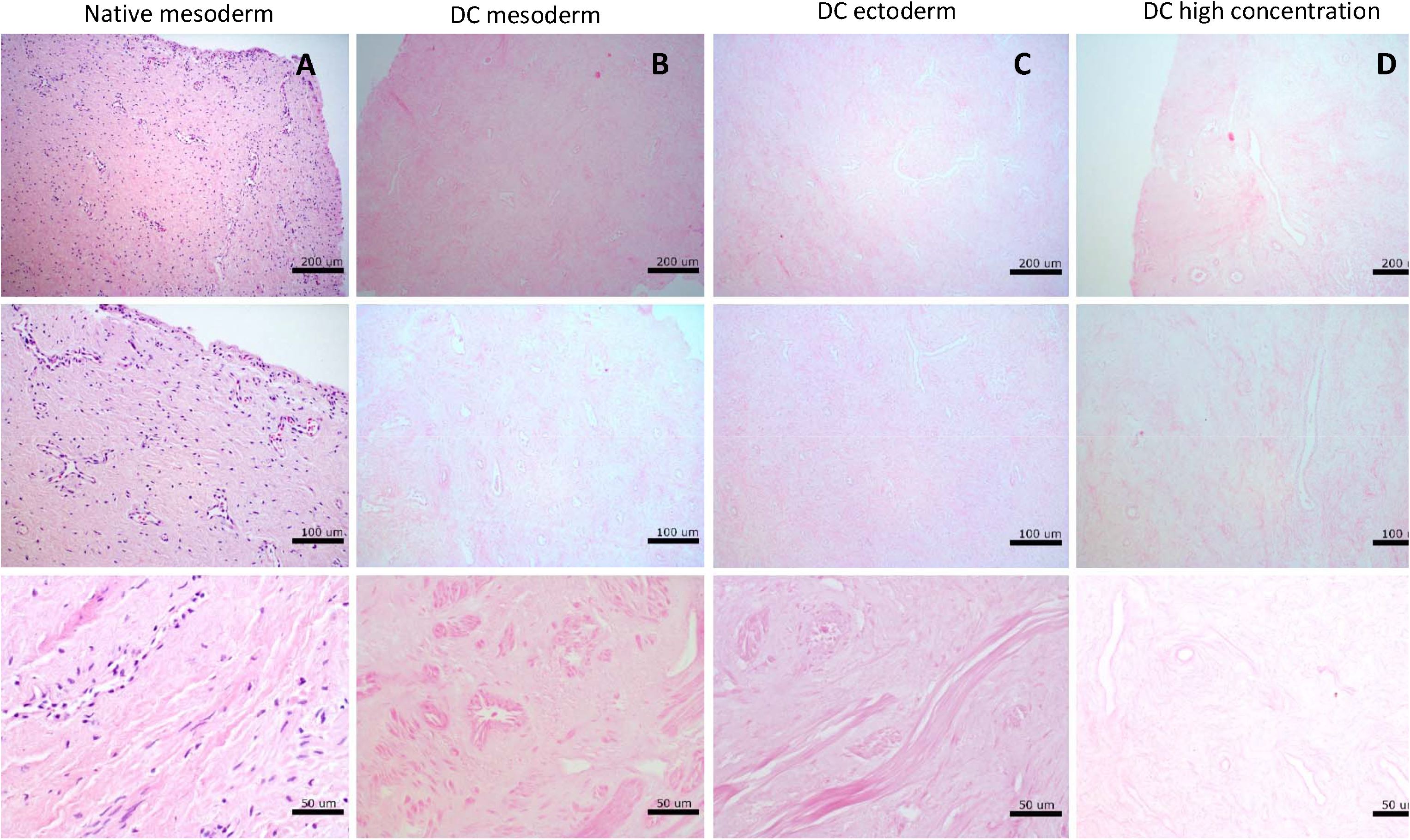
Assessment of tissue structure with Haematoxylin and Eosin (H&E) staining of A) native mesoderm, B) decellularized mesoderm, C) decellularized ectoderm and D) decellularized mesoderm vaginal tissue at high concentration.

### Significant DNA quantity reduction after DC

Quantitative DNA- and RNA-residue analysis confirmed removal of nuclear cellular material by DC. DC significantly (P<0,001) reduced dsDNA in vaginal tissue blocks below 50 ng and 10% dsDNA/mg dry ECM weight (Figure 4A). DsDNA was preserved (P>0,05) after DC by membrane disruption (Figure 4A) and RNA (0,001<P<0,01) after DC by enzymatic digestion (Figure 4C). This indicates that both enzymatic digestion and membrane disruption are essential to our DC protocol. RNA was not significantly reduced by addition of 3 U/mL RNase A (Figure 4C).

**Figure 4:**
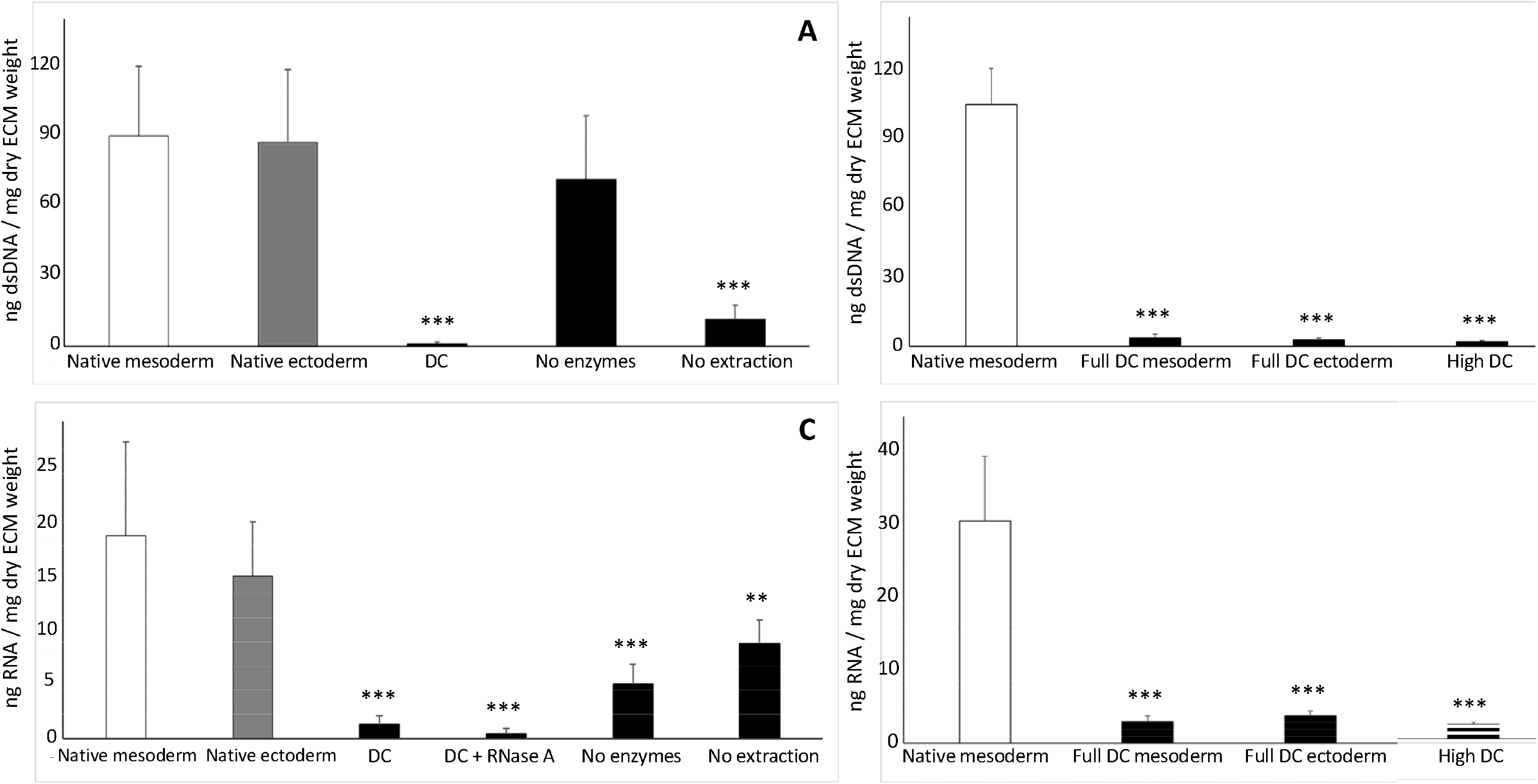
Quantification of residual dsDNA: A) Concentration of dsDNA is significantly (P<0,001) decreased in decellularized (DC) vaginal tissue and DC vaginal tissue without membrane disruption (“No extraction”) compared to native mesoderm and native ectoderm controls. B) Concentration of dsDNA is also significantly (P<0,001) decreased after DC of full intact vaginal tissue rings from Mesoderm and Ectoderm sections and for higher chemical concentrations during decellularization. Quantification of residual RNA: C) Concentration of RNA is significantly (P<0,001) decreased in decellularized (DC) vaginal tissue and DC vaginal tissue without enzymatic degradation (“No enzymes”) compared to native mesoderm and ectoderm conrols. D) Concentration of RNA is also significantly (P<0,001) decreased after DC of full intact vaginal tissue rings from Mesoderm and Ectoderm sections and for higher chemical concentrations during decellularization. The data represents the means of 7 experiments with standard deviation. Significance is depicted with *P<0,05, **P<0,01 and ***P<0,001.

In vaginal tissue rings, for clinically relevant sizes, dsDNA (Figure 4B) and RNA (Figure 4D) were significantly (P<0,001) reduced by DC of Mesoderm and Ectoderm tissue. DsDNA reduced below 50 ng and 10% dsDNA/mg dry ECM weight. Double chemical concentrations (0,36% w/w Triton x-100, 0,1% w/w sodium-deoxycholate and 300 U/mL DNase I) for DC did not significantly further reduce dsDNA or RNA.

### Significant DNA fragmentation after DC

DNA-residue fragments were assessed (TapeStation System) and reduced below 200 bp by DC (Supplement S1). DNA fragment lengths increased with reduced DNase I concentrations. Therefore, we applied 150 U/mL DNase I, although 100 U/mL DNase I is sufficient for fragmentation. Fragments above 200 bp were observed after DC by enzymatic digestion and below 200 pb After DC by membrane disruption.

### Decreased strain at rupture, tensile stress and elastic modulus after DC

Structural integrity after DC was assessed by mechanical testing of elongation until rupture. Force and tensile stress were low and similar for native and DC tissue until double the original length, where elastic wall components were stretched. Force and tensile strength increased with strain until strain at rupture ε (Figure 5A and 5C) at 1,8x elongation for native and 1,6x elongation for DC Mesoderm tissue, above which force and tensile stress decreased with strain. Visually, fibre bundle ruptures were observed. Despite quantitative differences, this pattern was observed in all patient samples. Beyond 2,6x elongation, force and tensile stress were similar for native and DC tissue. Validity of test conditions was confirmed by full vagina ring elongation (Figure 5B and 5D).

**Figure 5:**
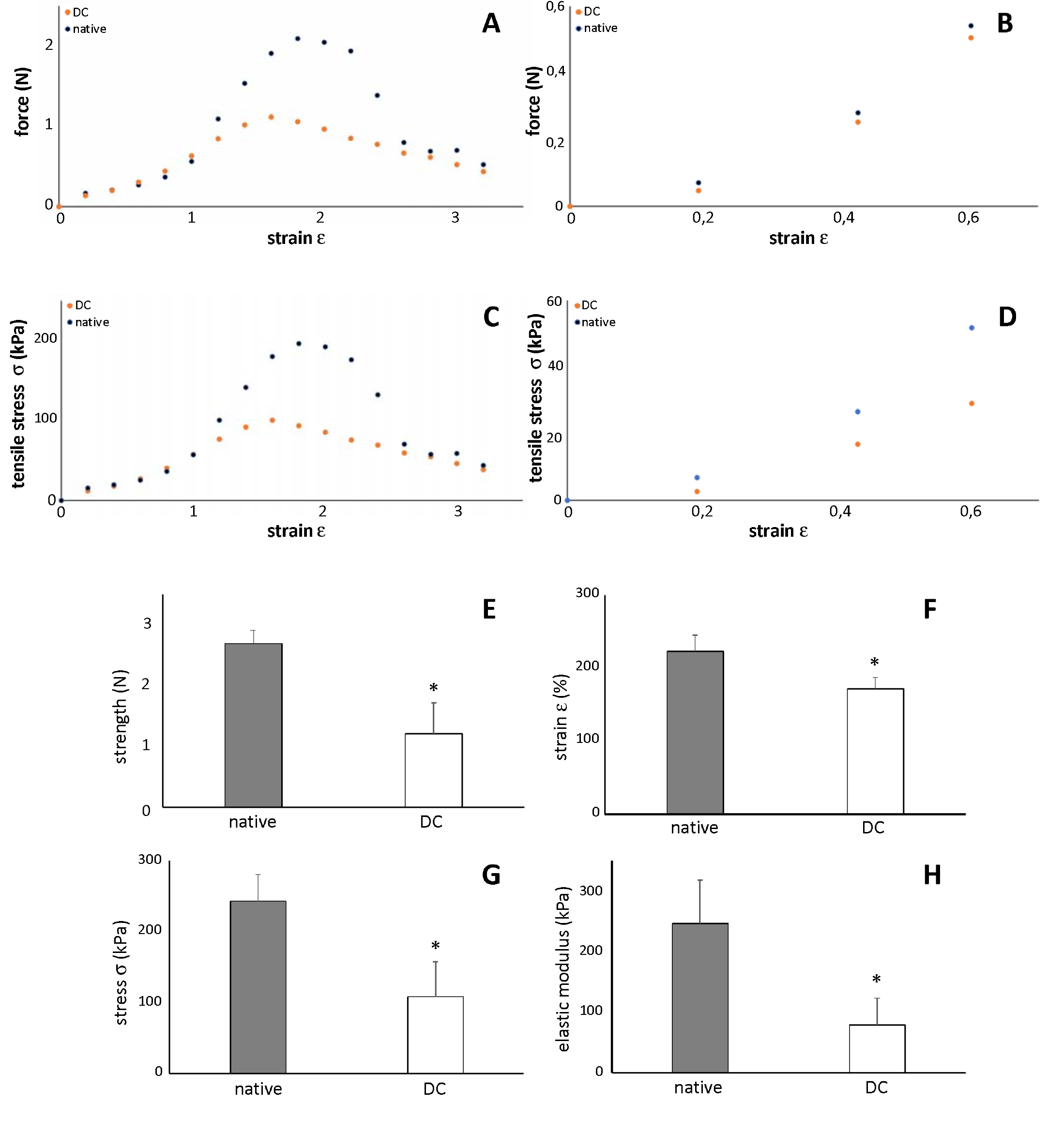
*Mechanical properties of human native and decellularized (DC) vaginal tissue*. Force as function of strain for (A) human vaginal tissue strips in Bioreactor and (B) human full vaginal rings in Ring stretcher. Tensile stress σ as function of strain for (C) human vaginal tissue strips in Bioreactor and (D) human full vaginal rings in Ring stretcher. The data represents the means of 7 experiments. *Mechanical properties of human native and decellularized (DC) vaginal tissue, measured with tissue strips in Bioreactor. (E) Pull strength. (F) Strain at rupture ε*. (G) Ultimate tensile stress σ. (H) Elastic modulus E. The data represents the means of 7 experiments with standard deviation. Significance is depicted with *P<0,05, **P<0,01 and ***P<0,001.

The pull strength (Figure 5E), strain at rupture ε (Figure 5F – strain at maximum stress), ultimate tensile stress σ (Figure 5G) and elastic modulus *E* (the resistance to elastic deformation) (Figure 5H) decreased by DC with significance (0,01<P<0,05). There was a decrease of 55% strength, 23% strain, 55% stress and 68% elastic modulus.

### Major constitutive ECM proteins retained after DC

Laminin-1/2 was mainly found around superior layers of the vaginal epithelium in native and DC vagina rings. Despite an immunofluorescent signal decrease, Laminin was mostly retained after DC (Figure 6). Fibronectin-I/II/III (Figure 7), Collagen-I (Figure 8) and the Ficolin-2B (Figure 9) matrix proteins were evenly dispersed throughout the ECM. Fibronectin was retained while Collagen-I and Ficolin-2B were (slightly) reduced by decellularization.

**Figure 6:**
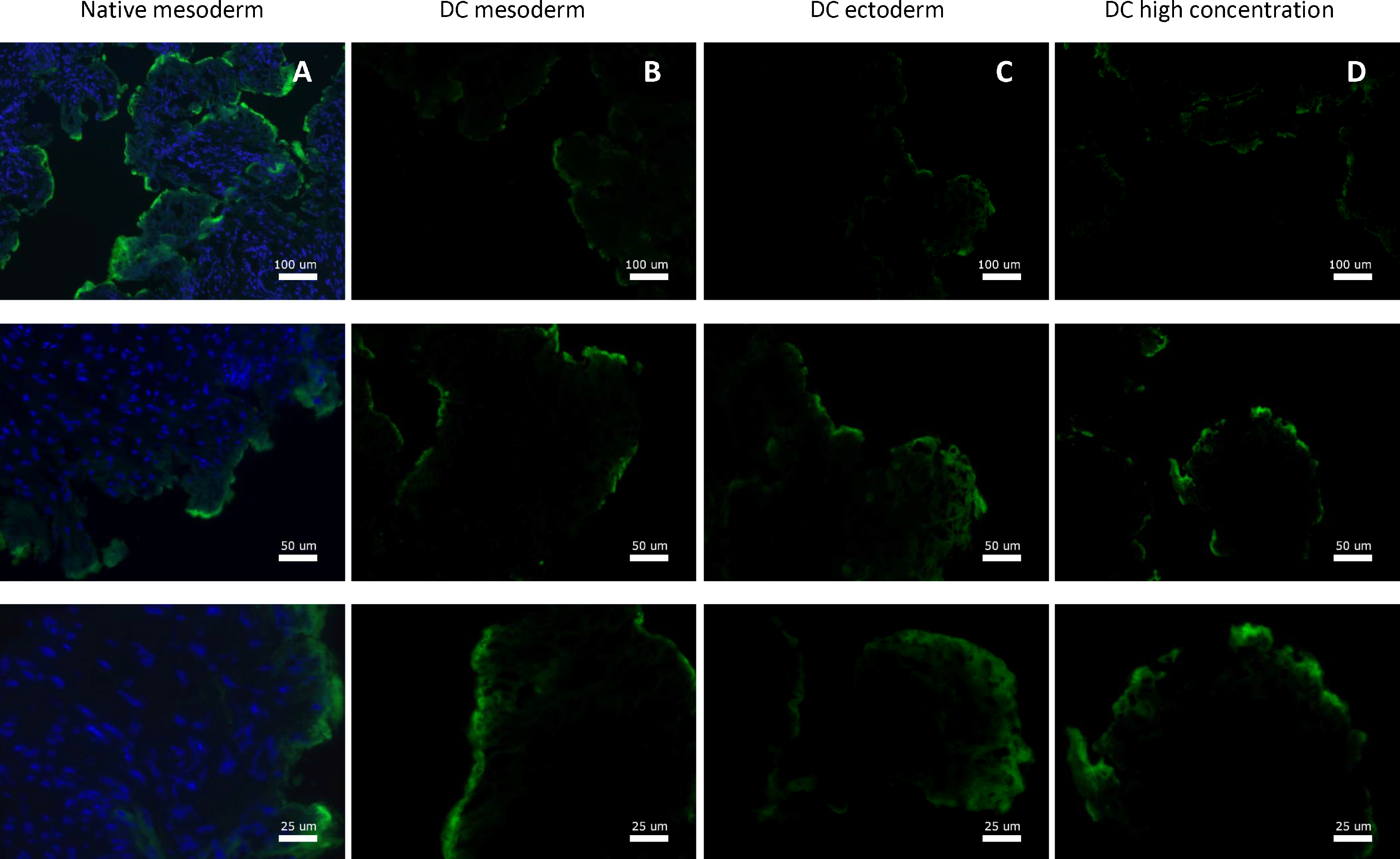
Fluorescence microscopy imaging of Laminin for vagina wall rings from A) native, B) decellularized mesoderm, C) decellularized ectoderm and D) high concentration decellularized mesoderm tissue. Fluorescence microscopy of Laminin was performed on and evaluated for all 7 patient samples.

**Figure 7:**
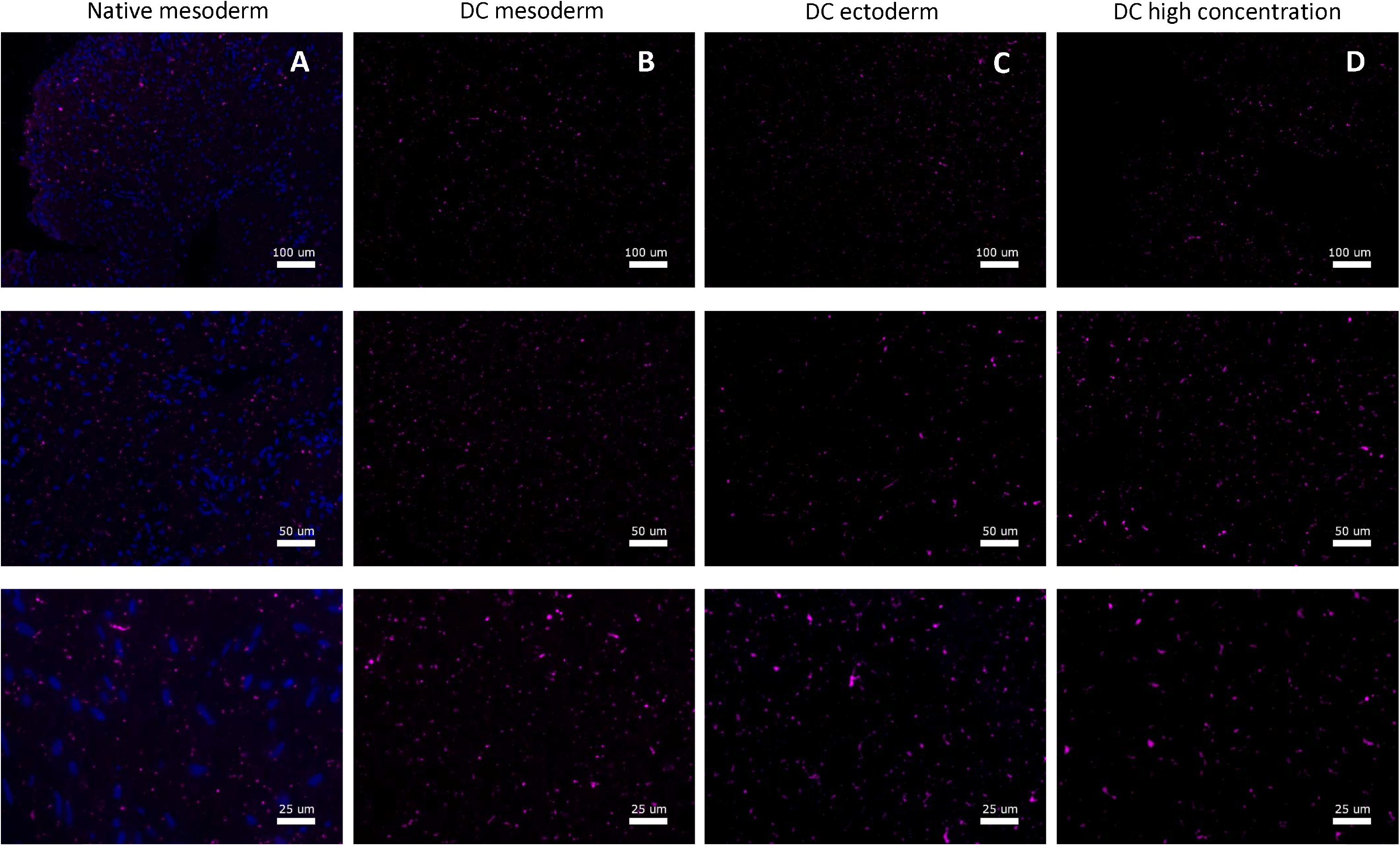
Fluorescence microscopy imaging of Fibronectin for vagina wall rings from A) native, B) decellularized mesoderm, C) decellularized ectoderm and D) high concentration decellularized mesoderm tissue. Fluorescence microscopy of Fibronectin was performed on and evaluated for all 7 patient samples.

**Figure 8:**
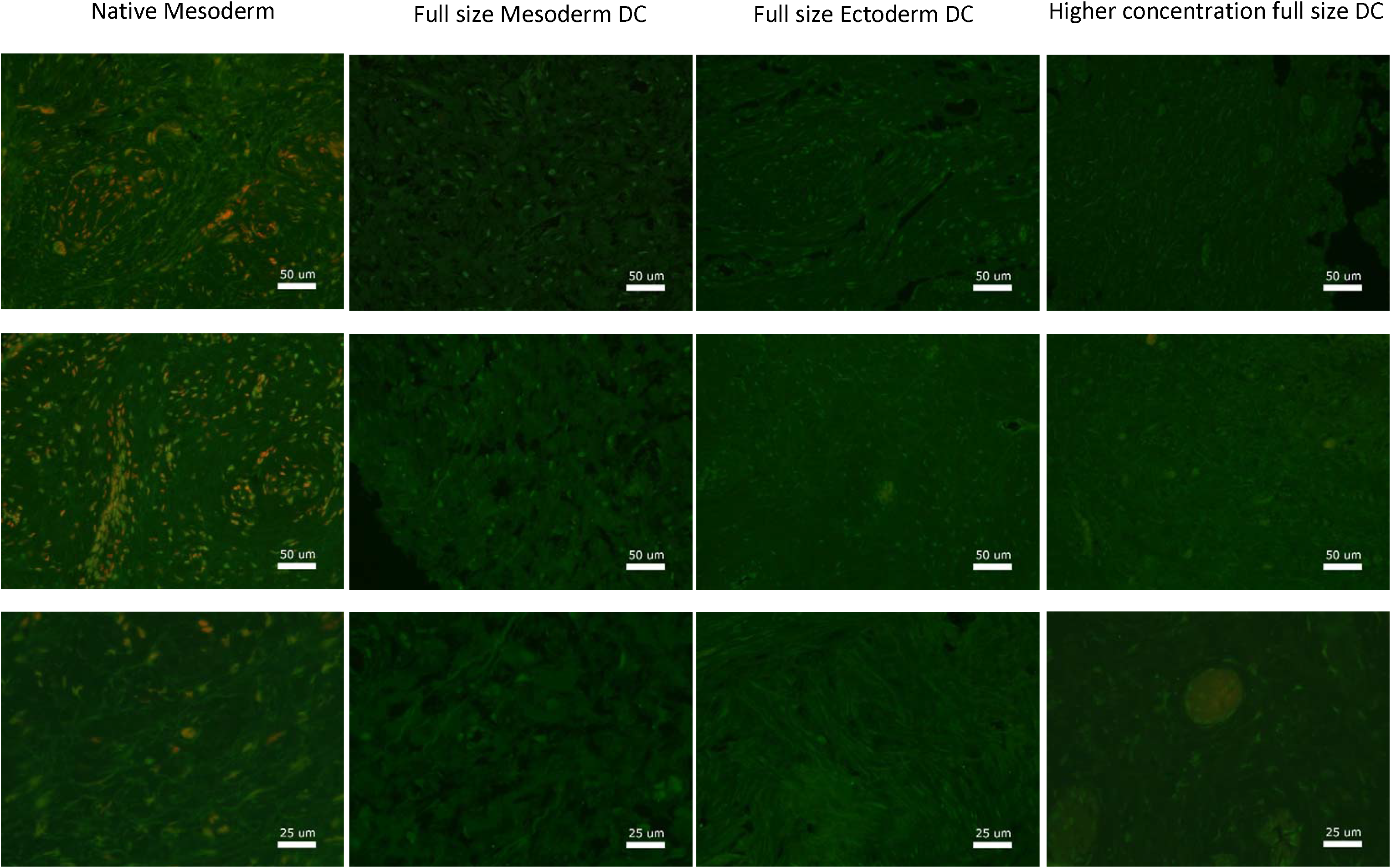
Fluorescence microscopy imaging of Collagen-I for vagina wall rings from A) Native, B) Decellularized Mesoderm, C) Decellularized Ectoderm and D) high concentration decellularized Mesoderm tissue. Fluorescence microscopy of Collagen-I was performed on and evaluated for all 7 patient samples.

**Figure 9:**
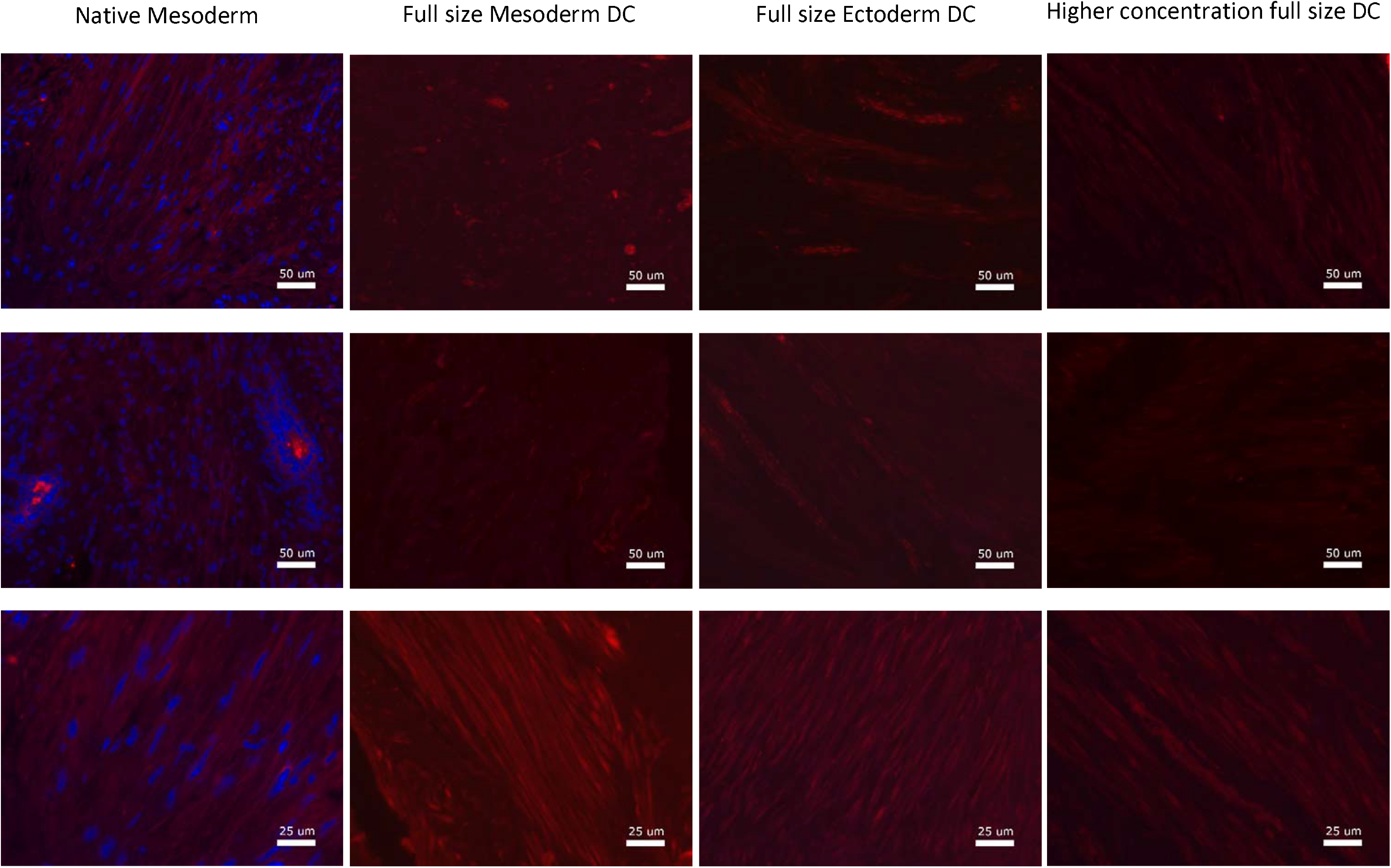
Fluorescence microscopy imaging of Ficolin for vagina wall rings from A) Native, B) Decellularized Mesoderm, C) Decellularized Ectoderm and D) high concentration decellularized Mesoderm tissue. Fluorescence microscopy of Ficolin was performed on and evaluated for all 7 patient samples.

## Discussion

### Results

Over 1.000.000 annual vaginoplasties are performed, but lack or absence of tissue, incompatible autologous grafts and immunorejection from allogenic grafts pose difficulties. Decellularization of vaginal matrix could prevent immunogenicity whilst preserving structural proteins and tissue structure for functionality and biocompatibility. Our study showed successful decellularization of human vaginal tissue based on absence of visible nuclei, dsDNA quantity <50 ng AND <10%/mg dry ECM weight and remnant DNA fragments <200 bp.

Visible nuclei in vaginal mesoderm and ectoderm tissue were fully removed after decellularization without chemically-induced loss of structural features in DAPI- and H&E-stained tissue. Both enzymatic degradation and membrane disruption are therein detrimental. Membrane disruption was incapable of satisfactional DNA and RNA removal, but enzymatic degradation caused structural alteration.

Decellularization significantly reduced (P<0,001) dsDNA to <50 ng AND <10%/mg dry ECM weight at <200 bp fragments, it also significantly reduced RNA quantity. Membrane disruption was incapable of successful DNA removal but significantly reduced RNA quantity with successful DNA fragmentation. Enzymatic degradation was incapable of successful DNA fragmentation but significantly reduced dsDNA quantity.

Successful decellularization of human vaginal matrix relied on both membrane disruption and enzymatic degradation. DNA mobility decreases with length and depends on immobilisation, random diffusion, active transport, anomalous subdiffusion, confined diffusion, transient confinement and binding-unbinding mechanisms [128], [129]. Short cytoplasmic DNA (<240 bp) is highly mobile through fast active transport and binding-unbinding [129], long DNA moves by slow anomalous subdiffusion and molecular crowding and nuclear DNA is almost immobile [128], [129]. Membrane disruption breaks the cellular membrane [123], thus removes mitochondrial DNA (and cytoplasmic RNA), but nuclear DNA requires enzymatic fragmentation for mobilization to allow diffusion. At the same time, DNA fragmentation by enzymatic degradation relies on membrane disruption (likely by improving accessibility) for success.

According to the commonly applied acellular criteria [130], decellularization of human vaginal tissue was successful. However, these criteria only assess cellular material contamination based on DNA. DNA differs from other intra- and extracellular components (including RNA, proteins, amino acids, lipids and Glycosaminoglycans) in physiology and chemistry and thus the interaction with decellularization agents [130]. In our opinion, assessment of decellularization should therefore always be supplemented with evaluation of tissue structure and function [132]. As criteria on these aspects are non-existent, we supplemented the definition of successful decellularization with: visible structural features, biocompatibility during stretching and presence of visible collagen, elastin, laminin and fibronectin. Our study also showed successful decellularization based on tissue structure and function, with space for improvement on biocompatibility and on preservation of Collagen-I and Ficolin-2B.

Our optimized protocol preserved vaginal tissue structures. Enzymatic degradation caused structural alterations by non-extracted, fragmented DNA and other cellular waste components that build up after a five-day DC protocol by incompletely removal. Double chemical concentrations induced structural changes. Many decellularization methods are known to cause structural damage [121], [123] and should be avoided. Our included donors were presurgically exposed to testosterone for mean=5,9 (range 2-11) years, with no vaginal atrophy (thinned squamous epithelium) or prostatic metaplasia (epithelial or subepithelial stroma glands) present in our samples. Induced structural alterations have been reported as prostatic metaplasia in 7 MtF-patients (18-34 years) after 24-72 months of presurgical exposure to testosterone cypionate and metastatic glands located predominantly in epithelial layers [133] or as 80% atrophy, 40% urothelial metaplasia and 70% prostate-like glands in 10 MtF-patients (22-74 years) after 2-10 years of preoperative androgen receival [134].

Laminin-I/II and Fibronectin-I/II/III were visibly retained and Collagen-I and Ficolin-2B were reduced after DC. Strength, stiffness and flexibility were slightly reduced (P<0,05) after DC. Native and decellularized tissue responded similar up to strain = 1 (elongated to twice the initial length), so under daily activity-like conditions. Loss of mechanical strength, can be explained by cavities, loss of osmotic pressure by cell removal from the matrix [135] and weakened matrix after removal of muscle cells and proposedly by damaged collagen crosslinks [136]. This correlates with our visibly reduced collagen and elastin. Tensile properties are inhibited by loss of proteoglycans or Fibronectin. Fibronectin-I/II/III were retained, but Proteoglycan are reportedly reduces by decellularization [85], [137], [138]. Strength and elasticity are dependent on expression of multiple extracellular matrix proteins, including collagen and elastin [139], [140]. The strength and elasticity decrease are therefore explained by the visible reduction of Collagen-I and Ficolin-2B.

### Impact of results

Our results confirmed that the multistep decellularization protocol is effective to prepare decellularized vaginal grafts from healthy full human donor tissue, with retained structure, sufficient biomechanical properties and (partial to full) conservation of structural proteins. These grafts are promising biomaterials for vaginal reconstruction, as the orthotopic origin should increase functionality by reduced complications that arise from nonvaginal tissue. Our DC protocol is successful for vaginal tissue, but is also suitable for any type of (human) tissue after optimization of chemical concentration. However, some future optimization of matrix functionality is vital through Collagen-I and Ficolin-2B restoration for required biomechanical properties, like stiffness and strength as a hollow organ, and flexibility during sex and in natural child birth.

## Future prospective

In future, our decellularization protocol will be supplemented with autologous host cell recolonization to test biocompatibility and cell impact on Collagen-I and Ficolin-2B restoration. Furthermore, incorporation of neovascularization for increased tissue survival and tissue innovation for biomechanical optimization are crucial aspects for successful vagina transplantation that need to be addressed.

## Data Availability

All data produced in the present study are available upon reasonable request to the authors.

## Acknowledgements

We would like to thank Remco Keijzer, Saskia van Daalen and Cindy de Winter-Korver for their assistance with new experimental protocols. We also like to share our gratitude towards Wendy Noort for the input and assistance with biomechanical testing.

## Authorship contribution statement

J.S.: Conceptualization, data curation, formal analysis, investigation, methodology, project administration, software, visualization, writing – original draft. F.X.: Data curation, investigation, methodology, software, visualization. F.G.: Resources, supervision, validation, writing – review & editing. H.M.: Conceptualization, investigation, methodology, resources, software, visualization. J.H.: Conceptualization, funding acquisition, methodology, resources, supervision, writing – review & editing. T.S.: Conceptualization, formal analysis, investigation, methodology, supervision, validation, visualization, writing – review & editing.

## Supplementary data

Supplementary data are available at *Biomaterials* online.

## Footnotes

1 MRKHS incidence is 1:1.500-10.000 female births [21], [141], [142], with 10-25% surgical treatments.

2 GD incidence is 1:2.900-45.000 genotypic males [143]–[147], with 5-38% surgical treatments [147]–[151]. Our Gender Clinic showed an incidence of 1:3800 [152] with 38% surgical treatment according to Dutch research [151].

3 DAPI (minor groove binder) has a high DNA affinity with high fluorescence intensity, whereas EtBr (intercalator) binding has low fluorescence with high RNA binding affinity.

